# Predicting Cost-Effectiveness of Generic Versus Brand Dabigatran Using Pharmacometric Estimates among Patients with Atrial Fibrillation in the United States

**DOI:** 10.1101/19006346

**Authors:** Ching-Yu Wang, Phuong N. Pham, Sarah Kim, Karthik Lingineni, Stephan Schmidt, Vakaramoko Diaby, Joshua Brown

## Abstract

Generic entry of newer anticoagulants is expected to decrease the costs of atrial fibrillation management. However, when making switches between brand and generic medications, bioequivalence failures are possible. The objectives of this study were to predict and compare the lifetime cost-effectiveness of brand dabigatran with hypothetical future generics. Markov micro-simulations were modified to predict the lifetime costs and quality-adjusted life years of patients on either brand or generic dabigatran from a U.S. private payer perspective. Event rates for generics were predicted using previously developed pharmacokinetic-pharmacodynamic models. The analyses showed that generic dabigatran with lower-than-brand systemic exposure was dominant. Meanwhile, generic dabigatran with extremely high systemic exposure was not cost-effective compared to the brand reference. Cost-effectiveness of generic medications cannot always be assumed as shown in this example. Combined use of pharmacometric and pharmacoeconomic models can assist in decision making between brand and generic pharmacotherapies.

## INTRODUCTION

Atrial Fibrillation (AF) is the most common cardiac arrhythmia caused by inconsistent muscle contraction of the atrium. AF increases the risk of systematic embolism, ischemic stroke and long-term mortality (1-3). Antithrombotic therapy with oral anticoagulants has been shown to lower the risk of stroke (4, 5). Patients with a high risk of stroke (e.g. CHA_2_DS_2_-VASc score ≥ 2) are recommended to receive long-term anticoagulation (6-9). Anticoagulation at the same time increases the risk of major bleeding including intracranial (ICH) and extracranial hemorrhage (ECH) (10). Thus, the risks versus benefits of anticoagulation and varying treatment options must be considered.

Warfarin, the long-standing mainstay of therapy, has been supplanted by direct oral anticoagulants (DOACs) as the treatments of choice for stroke prevention in AF. The first DOAC, dabigatran, (Pradaxa®) was approved by the Food and Drug Administration (FDA) in 2010. Compared to warfarin, dabigatran was shown to be non-inferior to warfarin with regards to the prevention of stroke or systemic embolism (11). Furthermore, in that study, major bleeding was less prevalent in the dabigatran treatment arm (11). Even though DOACs have been introduced to the market eight years ago and with favorable efficacy and safety profiles, the cost-effectiveness profiles of DOACs, particularly brand dabigatran, has no clearly been established due to increased treatment costs. The eventual market entry of generic DOACs may change market dynamics through increased utilization of cheaper generic DOACs. As it was first approved, dabigatran is also the first expected generic DOAC, estimated to enter the market in 2021, which should substantially decrease costs for stroke prevention in AF.

While the entry of generic dabigatran should be beneficial to both patients and payers through decreased costs, pharmaceutical and pharmacologic complexity can often raise concerns regarding bioequivalence (BE) versus the reference brand product (12). Under current regulations, generic manufacturers need to demonstrate BE to the reference product by conducting studies on healthy volunteers as a surrogate for therapeutic equivalence (13, 14). Two pharmacokinetic parameters are often measured in the BE studies — C_max_ (maximum observed concentration) representing the absorption rate, and AUC (Area under the plasma concentration-time curve) representing the extent of absorption (13, 14). Under US Food and Drug Administration guidance (15), BE can be declared when the 90% confidence interval for the ratio of the population geometric means of the pharmacokinetic measures (such as C_max_ and AUC) for generic to brand product falls within 80%-125% or F=0.80 and F=1.25 (Figure 1).

**Figure 1.**
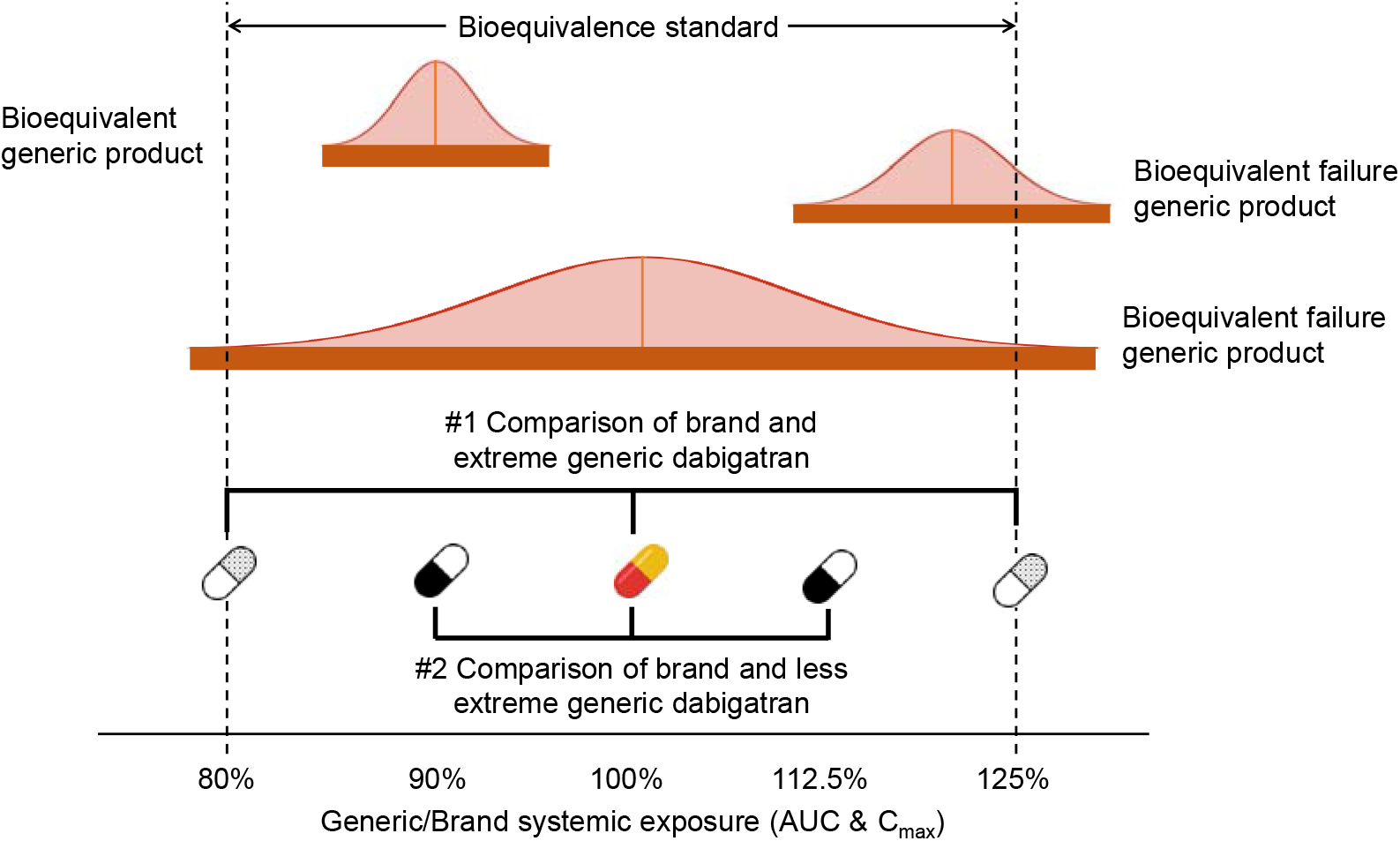
Demonstration of potential bioequivalent results and two comparisons made in this study. The solid orange bars represent the 90% confidence intervals of the bioequivalence study AUC and Cmax generic/brand ratios normally distributed around the geometric mean generic/brand ratio. Falling beyond 80-125% thresholds is a “ failure” of bioequivalence. Comparison #1 compares brand to generic dabigatran with extreme systemic exposure (125% and 80%). Comparison #2 compares brand to generic dabigatran with less extreme systemic exposure (112.5% and 90%). AUC= area under the plasma concentration-time curve; C_max_= maximum observed concentration.

The BE requirement for generic medications assumes that declarations of BE are consistent with therapeutic equivalence. However, due to batch-to-batch variability and between manufacturer differences, this may not always hold true. In fact, this phenomenon has been observed in examples of warfarin brand-to-generic comparisons. One observational study found international normalized ratios (INR) were significantly lower after switching from brand to generic warfarin(16). Another study found the number of visits with INR values outside of therapeutic range and the number of dose changes were higher in patients on generic warfarin(17). Switching from brand to generic warfarin was associated with a higher risk of thrombotic events (HR=1.81; 95% CI 1.42 to 2.31) and hemorrhagic events (HR=1.51; 95% CI 1.17 to 1.93)(18). Results from these studies may also call into question the clinical effects of generic medications and the economic benefits generic medications might bring.

These concerns are especially relevant to anticoagulants due to relatively narrow therapeutic indices and severity of both safety and effectiveness outcomes (i.e. hemorrhage and stroke). As dabigatran will be first to be offered as generic, these concerns raise questions regarding its known PK/PD profile, possible causes of variation in C_max_ and AUC, and how this will impact the entire therapeutic area. Incorporating PK/PD, clinical, and cost evaluations prior to generic entry may be informative to establish the value proposition of generic formulations, especially amid situations where some uncertainty in BE and therapeutic equivalence is predicted. The incorporation of pharmacometric (PK/PD) models that predict an exposure-response curve based on BE parameters for this application is novel and a unique application of these approaches.

Thus, the present study aimed to: (1) predict and compare the “ pre-launch” lifetime effectiveness of generic and brand dabigatran; (2) predict and compare the “ pre-launch” lifetime overall medical cost of generic and brand dabigatran; (3) evaluate the “ pre-launch” cost-effectiveness of generic dabigatran versus the reference brand in AF patients. This study expands upon base pharmacometric models to extrapolate parameters to clinical decision-making through cost-effectiveness models.

## RESULTS

### Base case analysis

In the comparison of brand and hypothetical extreme generic dabigatran, the base case analysis showed the F=0.8 generic dabigatran had the highest effectiveness (QALYs) among the three versions followed by the brand dabigatran (Table 2). While the occurrence of stroke was similar across different versions of dabigatran, more variability was observed with bleeding events which was consistent with the predicted bleed and stroke rates from the PK/PD models. The cohort on the F=1.25 generic had an excess of roughly 2,000 more (14,112 vs. 12,193; 15.7% increase) bleeding events than the brand dabigatran and roughly 3,500 more (14,112 vs. 10,604; 33.1% increase) bleeding events than the F=0.8 generic dabigatran (Table 2). The F=0.8 generic dabigatran had the lowest cost among the three dabigatran treatments followed by the F=1.25 generic. The F=0.8 generic had both higher QALY and lower costs; thus, it dominated the other treatment options. Direct comparison between the F=1.25 and brand dabigatran showed the brand to be more cost-effective than the F=1.25 generic with an ICER value of $36,483/QALY.

**Table 1.** Key model input parameters.

| Variable | Base Case | Range | Reference |
| --- | --- | --- | --- |
| Cost in 2017 monthly (US\$) | | | |
| Brand dabigatran | 296.23 | -- | (23, 25) |
| Generic dabigatran | 257.72 | -- | (23-25) |
| Rate of ischemic stroke on brand dabigatran for different patient subgroups (%/year) |  |  |  |
| Low stroke risk subgroup<br>(CHA2DS2-VASc score 2-3) | 0.721 | (0.569-0.930) | (IBM, 11) |
| Medium stroke risk subgroup<br>(CHA2DS2-VASc score 4) | 1.082 | (0.854-1.396) | (IBM, 11) |
| High stroke risk subgroup<br>(CHA2DS2-VASc score $\geq 5$) | 1.942 | (1.533-2.300) | (IBM, 11) |
| Rate of minor bleeding on brand dabigatran for different patient subgroup (%/year) |  |  |  |
| Low bleed risk subgroup<br>(HAS-BLED score 0-1) | 7.361 | (6.876-7.846) | (IBM, 11) |
| Medium bleed risk subgroup<br>(HAS-BLED score 2) | 9.183 | (8.577-9.788) | (IBM, 11) |
| High bleed risk subgroup<br>(HAS-BLED score $\geq 3$) | 13.151 | (12.029-14.018) | (IBM, 11) |
| Rate of ICH on brand dabigatran for different patient subgroup (%/year) |  |  |  |
| Low bleed risk subgroup<br>(HAS-BLED score 0-1) | 0.199 | (0.167-0.298) | (IBM, 11) |
| Medium bleed risk subgroup<br>(HAS-BLED score 2) | 0.248 | (0.167-0.372) | (IBM, 11) |
| High bleed risk subgroup<br>(HAS-BLED score $\geq 3$) | 0.355 | (0.239-0.532) | (IBM, 11) |
| Rate of ECH on brand dabigatran for different patient subgroup (%/year) |  |  |  |
| Low bleed risk subgroup<br>(HAS-BLED score 0-1) | 2.050 | (1.494-2.395) | (IBM, 11) |
| Medium bleed risk subgroup<br>(HAS-BLED score 2) | 2.557 | (1.864-2.9875) | (IBM, 11) |
| High bleed risk subgroup<br>(HAS-BLED score $\geq 3$) | 3.662 | (2.670-4.279) | (IBM, 11) |
| Efficacy and safety of F=1.25 generic dabigatran |  |  |  |
| HR for ischemic stroke, F=1.25 generic vs brand | 0.934 | NA | (26) |
| HR for bleeding event (minor bleeding, ICH, ECH), F=1.25 generic vs brand | 1.211 | NA | (26) |
| HR for MI, F=1.25 generic vs brand | 1.062 | NA | (27) |
| Efficacy and safety of F=0.8 generic dabigatran |  |  |  |
| HR for ischemic stroke, F=0.8 generic vs brand | 1.066 | NA | (26) |
| HR for bleeding (minor bleeding, ICH, ECH), F=0.8 generic vs brand | 0.842 | NA | (26) |
| HR for MI, F=0.8 generic vs brand | 0.951 | NA | (27) |
| Efficacy and safety of F=1.125 generic dabigatran |  |  |  |
| HR for ischemic stroke, F=1.125 generic dabigatran vs brand | 0.967 | NA | (26) |
| HR for bleeding (minor bleeding, ICH, ECH), F=1.125 dabigatran vs brand | 1.1055 | NA | (26) |
| HR for MI, F=1.125 generic vs brand | 1.031 | NA | (27) |
| Efficacy and safety of F=0.9 generic dabigatran |  |  |  |
| HR for ischemic stroke, F=0.9 generic dabigatran vs brand | 1.033 | NA | (26) |
| HR for bleeding (minor bleeding, ICH, ECH), F=0.9 generic dabigatran vs brand | 0.921 | NA | (26) |
| HR for MI, F=0.9 generic vs brand | 0.976 | NA | (27) |
| Abbreviations: F=bioavailability ratio versus reference brand; HR=hazard ratio; ICH=intracranial haemorrhage; ECH=extracranial haemorrhage; MI=Myocardial Infarction |  |  |  |

**Table 2.** Base Case Comparison between Brand and Hypothetical Extreme Generic Dabigatran Formulations.

| Treatment Strategy | Therapies in the Order of Cost |  |  |  |  | Dominance |
| --- | --- | --- | --- | --- | --- | --- |
|  | Cost (95% CI) | QALY (95% CI) | Stroke, N | All bleeding, N |  |  |
|  |  |  |  | Minor bleeding, N | Major bleeding, N |  |
| Generic dabigatran (F=0.8) | \$49,510 (\$7,319-\$195,785) | 7.38 (0.29-19.94) | 2,212 | 10,604 | | Dominant* |
|  |  |  |  | 7,615 | 2,989 |  |
| Generic dabigatran (F=1.25) | \$50,937 (\$7,319-\$206,008) | 7.29 (0.29-19.78) | 2,192 | 14,112 | | Absolutely dominated |
|  |  |  |  | 10,207 | 3,905 |  |
| Brand dabigatran | \$53126 (\$7,453-\$203,844) | 7.35 (0.29-19.85) | 2,174 | 12,193 | | Absolutely dominated |
|  |  |  |  | 8,785 | 3,408 |  |
| Abbreviations: F=bioavailability ratio versus reference brand; CI=confidence interval; QALY=quality-adjusted life years |  |  |  |  |  |  |
| *Dominant implies both lower costs and higher effectiveness (QALYs) |  |  |  |  |  |  |

The comparison of brand and less extreme hypothetical cases of generic dabigatran versions showed consistency in that the F=0.9 generic dabigatran was dominant with the highest effectiveness (QALYs) and the lowest cost (Table 3). The difference in the occurrence of clinical events was also less extreme. Similar to the other comparison, the occurrence of stroke event was almost the same across three dabigatran treatment groups. The cohort on the F=1.125 dabigatran had an excess of roughly 1,000 more (13,167 vs. 12,125; 8.6% increase) bleeding events than the brand dabigatran and nearly 1,800 more (13,167 vs. 11,391; 15.6%) bleeding events than the F=0.9 generic dabigatran (Table 3). Direct comparison between brand and F=1.125 generic dabigatran showed the brand retained more utility than the F=1.125 generic version but with a much higher ICER of $287,200/QALY compared to the more extreme generic (F=1.25).

**Table 3.**
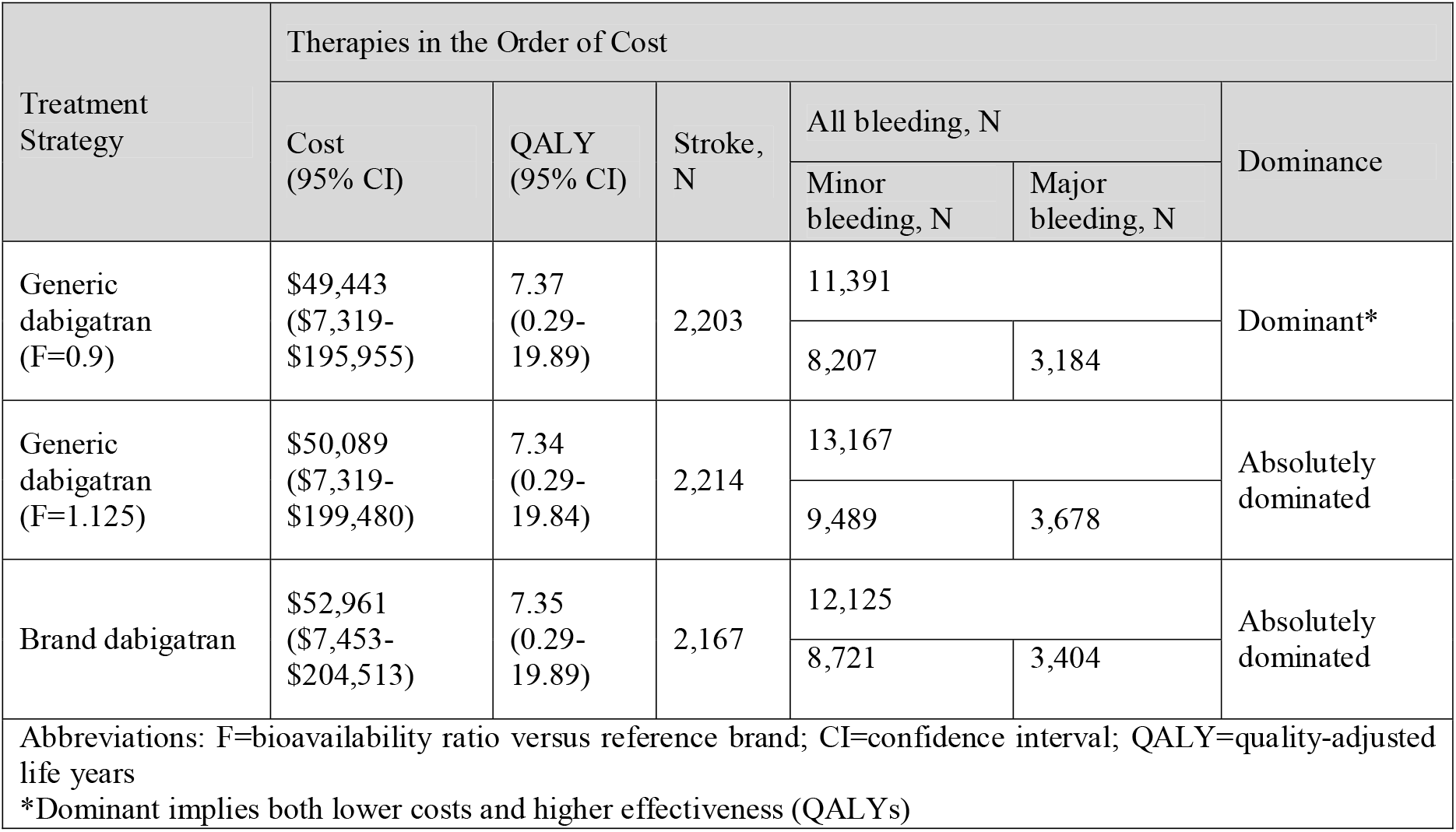
Comparison between Brand and Generic Dabigatran with Less Extreme Systemic Exposure.

### Sensitivity analyses

One-way sensitivity of the comparison between brand and extreme generic dabigatran versions showed the F=0.8 generic was the optimal treatment option under all tested conditions. While results from direct comparison between brand and F=1.25 dabigatran was robust to most of the parameter changes, F=1.25 generic was more cost-effective than the brand at a WTP threshold of $50,000/QALY in the following cases:

∘ Monthly cost of generic dabigatran lower than $248 (Figure 4A)

**Figure 2.**
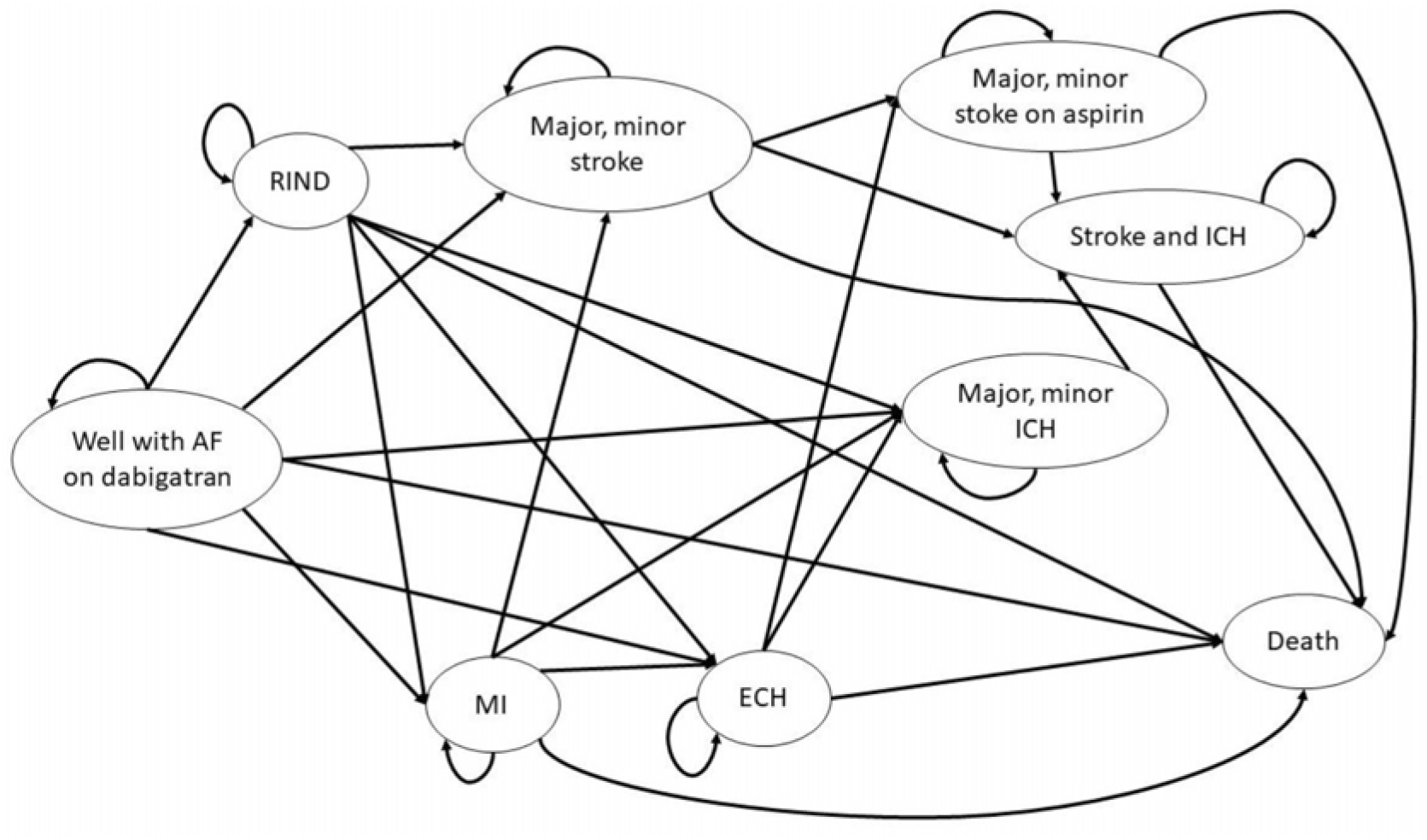
Diagram of the Markov model transition states. State transition diagram of Markov model shows all patients start with atrial fibrillation and then cycle between health states until death occurs. Major and minor stroke state were combined for demonstration purpose, so do the major and minor stroke on aspirin, as well as major and minor ICH. RIND=reversible ischemic neurological damage; ICH=intracranial hemorrhage; ECH=extracranial hemorrhage; MI=myocardial infarction.

**Figure 3.**
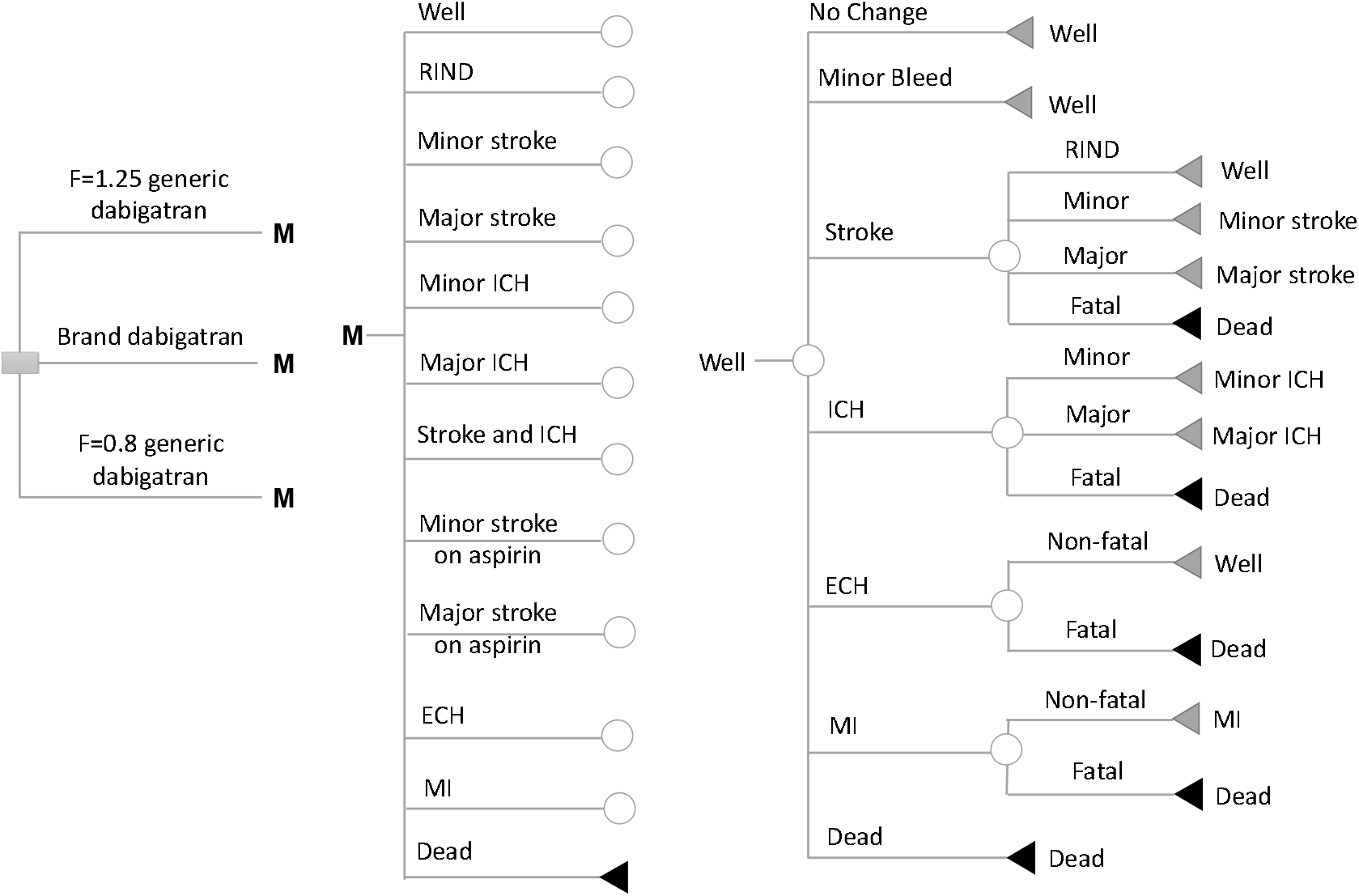
Schematic representation of Markov model shows the possible transitions for patients in well state. 10 other health states (except for death state) have similar structure of clinical event patients could encounter but different jump-to states. Probabilities of these events depend on prescribed medications and individual stroke/bleed risk. Decision node (square), chance node (circles), and terminal nodes (triangles) are depicted.

**Figure 4.**
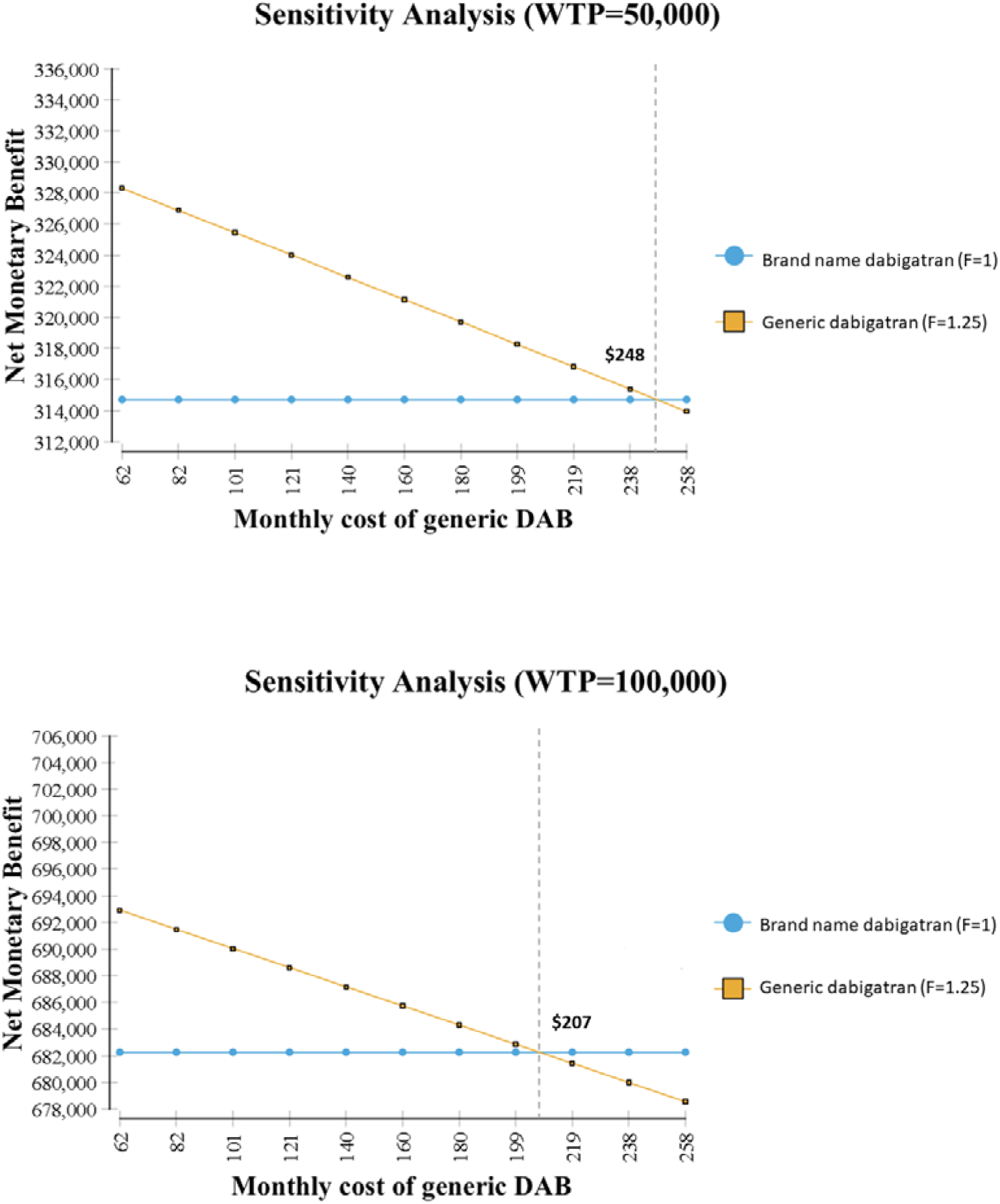
Results of one-way sensitivity analyses for cost of generic version. Net monetary benefit of brand and generic dabigatran with higher-than-brand extreme systemic exposure A) with a willingness-to-pay threshold of 50,000 B) with a willingness-to-pay threshold of 100,000.
∘ HR for ECH comparing dabigatran to warfarin was < 1.04
∘ MI rate for patients on warfarin < 1.06
∘ HR for MI comparing dabigatran to warfarin < 1.27 or > 1.85
∘ HR for ICH comparing dabigatran to warfarin < 0.33.

F=1.25 generic was also more cost-effective than the brand at a WTP threshold of $100,000/QALY in the following situation:

∘ Monthly cost of generic dabigatran lower than $207 (Figure 4B)
∘ HR for ECH comparing dabigatran to warfarin was < 0.91 (Supplementary Figure S7).

Probabilistic sensitivity analyses showed that the F=0.8 was the most cost-effective treatment option among the three in 100% of the iterations (Supplementary Figure 15).

One-way sensitivity of the comparison between brand and less extreme generic dabigatran showed F=0.9 generic was the optimal treatment option under all tested conditions (Supplementary Figure S8-S11). Results from the direct comparison between brand and F=1.125 dabigatran were robust to most of the tested conditions (Supplementary Figures S12-S14). Probabilistic sensitivity analyses showed that the F=0.9 was the most cost-effective option among the three in 99% to 100% of the iterations depending on the WTP threshold used (Supplementary Figure S16).

## DISCUSSION

The objective of this study was to compare the cost-effectiveness of hypothetical generic versions of dabigatran versus the brand. Two comparisons were conducted based on two assumptions of generic dabigatran systemic exposure—extreme cases (F=0.8 and F=1.25) and less extreme cases (F=0.9 and F=1.125). Our model shows that the generic dabigatran with the lower-than-brand systemic exposures (F=0.8 and F=0.9) were the most cost-effective options in both comparisons with the highest QALY and lowest cost. However, as expected, the QALY difference between F=0.8 generic and brand was a minimal 11 days. The difference between F=0.9 and brand was an even smaller 7 days. The results are robust in that in the base case analysis and across all sensitivity analyses including probabilistic sensitivity analysis, F=0.8 and F=0.9 generic dabigatran were the optimal treatment strategies.

When the dominant treatment strategies (F=0.8 and F=0.9 generic dabigatran) were excluded, and only F=1.25 and F=1.125 generic dabigatran were compared with the brand, our model showed different results in two comparisons. In the extreme case, brand was more cost-effective than the F=1.25 generic, while in the less extreme case, F=1.125 generic was a preferred choice than the brand. The discrepancy between two comparisons could be explained by the smaller QALY difference in the less extreme case (4 days) than the QALY difference in the extreme case (22 days). The results are robust to most tested conditions.

Note that the price of generic dabigatran was estimated from a previous study (19) under the assumption that there is only one generic dabigatran available (generic costs 87% of the brand). The same study also reported the generic to brand price ratio when there are multiple generic medications available. Based on their estimations, when there are two generic dabigatran on the market, the generic dabigatran would have a price of $228 USD (i.e. 77% of the brand price). Under this scenario, the brand will no longer be cost-effective than F=1.25 generic dabigatran brand using a WTP threshold of $50,000/QALY but remained cost-effective using a WTP threshold of $100,000/QALY. However, when there are three generic dabigatran available, generic dabigatran would have an estimated price of $178 USD (the generic costs 60% of the brand). Under that scenario, the brand will no longer be cost-effective than the F=1.25 generic dabigatran using either $50,000 or $100,000 WTP threshold. Thus, these results, and the cost-effectiveness of generic dabigatran in extreme cases of increased bioavailability, may depend on competition and pricing of generics.

Anticoagulation as a therapeutic area has experienced past concerns about BE failures when generic versions of warfarin became available. Studies comparing brand and generic warfarin found decreased anticoagulation control, increased healthcare visits required to manage anticoagulation, and increased thrombotic and haemorrhagic events (16-18). Concerns with warfarin were due to its narrow therapeutic index – a concern that is lessened with DOACs. However, DOACs still have PK/PD profiles that can be of concern, particularly for bleeding events, which drove the major findings in this study. The choice of dabigatran as an example of combining pharmacometric parameters with CEA was motivated by convenience and projections of it being the first generic DOAC. This study does not necessarily imply that dabigatran has complex pharmaceutical or pharmacological properties that may lead to BE concerns or predict future product failures. However, dabigatran does have low bioavailability and is susceptible to drug-drug interactions. Paired with a relatively steep bleed risk curve with increasing bioavailability, the results of this study indicate that an increase in bioavailability of dabigatran, whether caused by formulation issues or drug-drug interactions, can lead to excess bleed events and lack of overall cost-effectiveness.

To the best of our knowledge, this is the first study to evaluate the cost-effectiveness of future, hypothetical generic formulations versus the brand using pharmacometrically derived input parameters (20). The results have important clinical implications. Such an approach, and the evidence generated, could enhance evaluation of generics by patients, providers, and healthcare payers with regards to generic medications. Combined, PK/PD models and pharmacoeconomic models can also be informative for regulatory bodies for the required evidence for generic approval (20). From a clinical standpoint, this study highlighted the clinical burden when the active ingredient of generic dabigatran was above that of the brand and demonstrated it was less of the concern when active ingredient of generic dabigatran was below that of the brand. Future studies which evaluate the cost-effectiveness of other generic medications with more complex pharmaceutical properties may be informative to healthcare providers and decision makers.

### Limitations

These results should be interpreted in light of a number of limitations. Effectiveness and safety estimates were obtained from clinical trials and further parameterized with external information from PK/PD models and may not be generalizable to real-world populations. Transition probabilities were further influenced by external information. To account for indirect evidence, we incorporated meaningful ranges and thorough one-way and probabilistic sensitivity analyses to observe the robustness of results amid this uncertainty. Like past cost-effectiveness models in AF, the model incorporates a time span much longer than the source material and does not incorporate additional incident events such as diabetes which would influence stroke and bleed risk scores. However, these events can generally be considered to be similar between treatment groups.

### Conclusion

Predictions of cost-effectiveness for future generic dabigatran products showed that lower-bound generics (F=0.80 bioavailability versus brand) was consistently cost-effective. Upper-bound generics (F=1.25 bioavailability versus brand) were not cost-effective, mainly due to an excess in bleeding events, though these results were sensitive to the cost of the generic version. Future generic DOACs should be more closely evaluated for sensitivity in bioavailability related to formulations and pharmaceutical parameters as these differences can lead to poor patient outcomes. Extensions of pharmacometric PK/PD models to cost-effectiveness and decision analyses can be informative for multiple stakeholders and provide insight into current or future value of generic medications.

## METHODS

### Economic model overview

A Markov model was adapted from Shah *et al*. to model the disease progression of AF (Figure 2 and 3) (21). We relaxed the cohort assumption inherent with Markov models to allow variations in patient clinical characteristics such as stroke and bleed risks and age. The model had a cycle length of 30 days with a lifetime horizon. The simulation was conducted from the U.S. private payer perspective. All state rewards including probabilities, costs, and health utilities were converted to reflect the 30-day cycle length. The half-cycle correction (HCC) was applied to adjust for the potential overestimation of the costs and utilities. The study population represented commercially insured adult patients with AF who are eligible to receive anticoagulants. The model population consisted of 10,000 AF patients aged 18 or above recommended for anticoagulation with a CHA_2_DS_2_-VASc score of 2 or above(22) and any value of HAS-BLED score(8, 22).

We updated the prior model to expand granularity for stroke and bleed risk estimates. We replaced the CHADS_2_ score with CHA_2_DS_2_-VASc score which has been shown to perform better in risk stratification(23, 24). We also incorporated individual bleed risk using the HAS-BLED score(25). Patients were grouped into nine exclusive subgroups based on the cross-tabulation of their stroke risk categories (low, moderate or high) and bleed risk categories (low, moderate, high). The joint distribution of CHA_2_DS_2_-VASc and HAS-BLED scores, as well as the age distribution for the nine subgroups, were derived from IBM MarketScan^®^ Databases, which was consistent with the commercial payer perspective of the model.

Clinical outcomes considered in the model were minor bleeding, stroke, ICH, myocardial infarction (MI), ECH, and death. The model structure consisted of twelve mutually exclusive health states: (1) Well with AF; (2) Reversible ischemic neurological damage (RIND); (3) Minor stroke; (4) Major stroke; (5) Major ICH; (6) Minor intracranial ICH; (7) Minor stroke on aspirin; (8) Major stroke on aspirin; (9) Stroke and ICH; (10) Myocardial infarction (MI); (11) Extracranial hemorrhage (ECH) and (12) death. Patients entered the model in the “ Well with AF” state on either brand or generic dabigatran and then transitioned to other health states based on the event transition probabilities. The following assumptions were made to reflect the actual treatment pattern of AF patients and to better approximate the disease progression of AF patients:

- 28% of all the ischemic strokes were transient ischemic attacks, the remaining could be 1 of 4 types: reversible, major, minor, or fatal;
- ICH could be of 3 types: major, minor, and fatal. ECH could be either fatal or non-fatal. After experiencing ECH or ICH, patients were assumed to discontinue dabigatran and switch to aspirin for the remainder of their life;
- After experiencing two major neurological events (stroke or ICH), patients will proceed to the death state;
- After experiencing two minor neurological events, patients will proceed to a major neurological event state.

For example, if patients in “ major ICH” state experience a major stroke, instead of entering “ major stroke” state, they will enter “ death” state. If patients in “ minor ICH” state experience another minor ICH, they will enter “ major ICH” state instead of staying in “ minor ICH state”. To estimate the net monetary benefits, we used two different willingness-to-pay (WTP) threshold of $50,000 and $100,000 per quality-adjusted life year (QALY). Costs and utilities were discounted at a rate of 3%.

Two sets of comparisons were performed. First, we compared brand dabigatran (F=1.0) with two hypothetical versions of generic dabigatran with extremes in systemic exposure based on AUC and Cmax. The two extreme generic dabigatran versions had, respectively, 80% systemic exposure of the brand (extreme lower-bound generic, F=0.8) and 125% systemic exposure of the brand (extreme higher-bound generic, F=1.25). Second, we compared brand dabigatran with two additional hypothetical dabigatran versions with less extreme values of BE, 90% and 112.5% (F=0.9 and 1.125, respectively). Figure 1 shows the brand to generic ratio of systematic exposure for the hypothetical generic dabigatran we compared in this study.

### Model input parameters

Key model input parameters are provided in Table 1 and a complete list of all input parameters, distributions, and data sources are provided in Supplementary Table S1. All costs were considered from a private payer’s perspective and only direct medical costs were included. The event costs of ICH, MI, stroke, and ECH and the follow-up costs of ICH, MI, and stroke were derived from AF-specific event and follow-up cost estimates. The cost for the branded dabigatran was estimated using the 2017 National Average Drug Acquisition cost compiled by the US Medicaid program(26). The cost for the generic dabigatran was calculated by multiplying the price for brand dabigatran by the reported relative cost difference between branded and generic drugs when only one generic drug is available(19). To more accurately reflect a payers’ perspective in which negotiated rebates are common when there are multiple branded products in a therapeutic category, we applied an average rebate of 23% to the price for both brand and generic dabigatran. All event and follow-up costs were inflated using the medical component of Personal Consumption Expenditures Price Index (PCEPI) to 2017 US dollars (27). The cost for aspirin was inflated using the pharmaceutical product component of PCEPI.

Utilities were accounted in each state and with each event to estimate cumulative QALYs for each patient. State utilities (state rewards) were attached to the 12 health states. Major and minor neurological events (stroke or ICH) were associated with a permanent annual disutility of -0.61 and -0.24, respectively. The decrements in utilities (transition rewards) were calculated for the transitional events (minor bleed, RIND, ECH and MI) in the model. MI, ECH and minor bleed were assigned temporary disutility of 30 days, 2 weeks, and 2 days, respectively. The detailed calculation of state utilities for the 12 health states and the transition utilities for the transitional events are shown in Supplementary Table S2.

The ischemic stroke and bleeding rates (minor bleeding, ICH, and ECH) for patients on brand dabigatran within each CHA_2_DS_2_-VASc score category were derived from IBM MarketScan^®^ Databases and the RE-LY trial(11). The difference in event rates (stroke and bleeding) between generic and brand dabigatran were obtained from the PK/PD model developed by Kim et al. by calculating a relative event rate for each bioavailability level (e.g. F=0.8 and F=1.25)(28). Relative risks for increased stroke and ICH with every decade were based on pooled analysis from clinical trials and systematic reviews. The MI rate for patients on brand dabigatran was obtained from the clinical trial. The difference between brand and generic in MI rate was approximated from a meta-analysis comparing 150 mg dabigatran to 110 mg dabigatran, which assumed a linear relationship between dabigatran dose and occurrence of MI(29). The calculations of the event rates and the data analysis were conducted in IBM MarketScan^®^ Databases and are described in detail in the supplementary materials.

Two cost-effectiveness analyses (CEAs) were performed to compare extreme generics and more BE generics each to the brand dabigatran. We performed a series of one-way sensitivity analysis by varying (1) cost of generic dabigatran; (2) the event and monthly costs; (3) the utilities; (4) the transition probabilities. The parameter ranges for one-way sensitivity analysis are shown in supplementary table S1. Probabilistic sensitivity analysis (PSA) was conducted to account for parameter uncertainty. The probability distribution reflected the nature of the data. When the simulation was initiated, a value for each input was randomly selected from its respective probability distribution. The model was run repeatedly for 100 iterations and the pooled results were summarized. Model development, implementation, and analyses were performed using TreeAge Pro 2018 (TreeAge Software Inc., Williamstown, MA, USA).

## Data Availability

All data (model inputs) are reported in the manuscript

## STUDY HIGHLIGHTS

### What is the current knowledge on the topic

- Generic versions of medications undergo bioequivalence testing to ensure similar bioavailability and similar therapeutic effects. Failures may occur when medications have complex pharmaceutical or pharmacological properties. Projecting the impact of variations in pharmacokinetics/pharmacodynamics and incorporating these parameters into cost-effectiveness models could inform clinical decision-making.

### What question did this study address

- This study considered whether or not generic versions are always cost-effective in light of bioinequivalence concerns. These analyses modelled hypothetical dabigatran generic versions using extreme values of bioequivalence thresholds and predicted event probabilities.

### What does this study add to our knowledge

- With dabigatran as an example, this study showed that generic medications are not always cost-effective when bioequivalence thresholds are considered. While the generic dabigatran with less extreme systemic exposure is cost-effective compared to the brand reference, generic dabigatran with extremely high systemic exposure is not cost-effective compared to the brand reference.

### How might this change clinical pharmacology or translational science

- This study implies that generic medications with complex pharmaceutical or pharmacological properties should be more closely scrutinized, not only by regulatory bodies, but also once on the market by prescribers and patients. This study also is indicative of the value of multidisciplinary and translational science to inform pharmacotherapy through the combination of pharmacometric and pharmacoeconomic approaches.

## ACKNOWLEDGEMENTS

*None*.

## AUTHOR CONTRIBUTIONS

C.W. and J.B. wrote the manuscript; C.W. and J.B. designed the research; J.B., S.S. V.D. provided intellectual input; C.W., P.P., S.K., K.L., S.S., V.D. and J.B. performed the research; C.W. and P.P. analyzed the data.

## SUPPLEMENTARY MATERIALS

Table S1. Complete list of input parameters, their distributions and data source

Table S2. The calculation of state and transition utilities

Detailed Calculation of Transition Probabilities (Event Rate)

IBM MarketScan® Database Analysis

Figure S1. Net monetary benefit of brand and extreme cases of generic dabigatran at varying generic dabigatran cost

Figure S2. Tornado diagram of net monetary benefit at varying event and monthly cost comparing brand and extreme cases of generic dabigatran

Figure S3. Tornado diagram of net monetary benefit at varying utilities comparing brand and extreme cases of generic dabigatran

Figure S4. Tornado diagram of net monetary benefit at varying transition probabilities comparing brand and extreme cases of generic dabigatran

Figure S5. Tornado diagram of incremental cost-effectiveness ratio comparing brand and F=1.25 generic dabigatran at varying event and monthly

Figure S6. Tornado diagram of incremental cost-effectiveness ratio comparing brand and F=1.25 generic dabigatran at varying utilities

Figure S7. Tornado diagram of incremental cost-effectiveness ratio comparing brand and F=1.25 generic dabigatran at varying transition probabilities

Figure S8. Net monetary benefit of brand and less extreme cases of generic dabigatran at varying generic dabigatran cost

Figure S9. Tornado diagram of net monetary benefit comparing brand and less extreme cases of generic dabigatran at varying event and monthly cost

Figure S10. Tornado diagram of net monetary benefit comparing brand and less extreme cases of generic dabigatran at varying utilities

Figure S11. Tornado diagram of net monetary benefit comparing brand and less extreme cases of generic dabigatran at varying transition probabilities

Figure S12. Net monetary benefit at varying cost of generic dabigatran comparing brand dabigatran and F=1.125 generic dabigatran

Figure S13. Tornado diagram of incremental cost-effectiveness ratio at varying event and monthly comparing brand dabigatran and F=1.125 generic dabigatran

Figure S14. Tornado diagram of incremental cost-effectiveness ratio at varying utilities comparing brand dabigatran and F=1.125 generic dabigatran

Figure S15. Results of the probability sensitivity analysis. Cost-effectiveness (CE) acceptability curve (probability that a treatment will be cost-effective at varying willingness-to-pay thresholds)

Figure S16. Results of the probability sensitivity analysis. Cost-effectiveness (CE) acceptability curve (probability that a treatment will be cost-effective at varying willingness-to-pay thresholds)

